# Causes of excess deaths in the US relative to other wealthy nations, 1999-2020: a population autopsy

**DOI:** 10.1101/2024.07.26.24311071

**Authors:** Jacob Bor, Rafeya Raquib, David Himmelstein, Steffie Woolhandler, Andrew C. Stokes

## Abstract

**Importance:** The US has higher mortality rates than other wealthy nations.

**Objective:** To determine causes of death responsible for excess mortality in the US compared to other wealthy nations and how the causes involved in this survival gap have changed over time.

**Design:** Repeat cross-sectional study, 1999 to 2020.

**Setting:** United States and 12 other wealthy nations.

**Participants:** All residents.

**Exposures:** Residing in the US versus other wealthy nations.

**Main outcome and measures:** Excess US mortality in each year due to specific causes of death using data from the World Health Organization Mortality Database. Differences between the US and other wealthy nations were quantified for each cause of death as: (1) the number of excess US deaths (i.e., deaths that would have been averted if US death rates equaled the average rates of other wealthy nations); (2) years of life lost (YLL) resulting from excess US deaths; and (3) the ratio of observed deaths to expected deaths if US mortality rates equaled the average of other wealthy nations.

**Results:** 10,856,851 excess US deaths occurred between 1999 and 2020. In 2019, prior to the COVID-19 pandemic, there were 637,682 excess US deaths, with leading causes including circulatory diseases (41% of total), mental and nervous system disorders (25%), diabetes, renal, and metabolic diseases (15%), drug poisonings, alcohol-related deaths, and suicide (13%), respiratory disease (12%), and transportation accidents (5%). Over two decades, excess US deaths due to drug poisonings, alcohol and suicide increased from -5,937 in 1999 to 109,015 in 2020. In 2019, deaths from drug poisonings were 6.7 times higher in the US than in peer countries. Circulatory mortality accounted for the largest absolute number of excess US deaths in nearly every year. In 2020, one in 5 excess US deaths were attributed to COVID-19.

**Conclusions and Relevance:** The US had substantially higher death rates than other wealthy nations between 1999 and 2020, despite having similar access to advanced medical technology. Many of these excess US deaths could likely be avoided by adopting health and social policies that have benefited peer countries.

**KEY POINTS:** *Question:* What causes of death are responsible for the survival gap between the US and other wealthy nations?

*Findings:* Between 1999 and 2020, 10,856,851 US deaths would have been averted if the US had mortality rates equal to the average of peer countries. Circulatory diseases were the leading cause of excess deaths, although deaths due to drugs, alcohol, and suicide increased the most during the study period. Mental and nervous system disorders, diabetes, renal, and metabolic diseases, and transportation accidents were also major contributors.

*Meaning:* The causes of death responsible for the US survival gap suggest areas for policy intervention.

## INTRODUCTION

US life expectancy has diverged from other wealthy nations since 1980, falling to 58th in global rankings in 2021.^1^ Between 1980 and 2021, 13 million US deaths – including 1.1. million in 2021 alone – could have been averted had US age-specific mortality rates equaled those of other wealthy nations. We have referred to these statistical excess US deaths as “Missing Americans.”^2^

Quantifying the causes of death responsible for excess US mortality may help illuminate avenues for prevention. Prior studies have focused on specific years,^3,4^ causes of death,^5–7^ or age groups.^8^ However, none has offered a comprehensive analysis encompassing all leading causes of death over the last two decades and extending into the COVID-19 pandemic.

To address this gap, we performed a “population autopsy” of the missing Americans, drawing on cause of death data for all deaths occurring from 1999 to 2020 in the US and 12 other wealthy nations.

## METHODS

### Data sources

We obtained raw counts of deaths by underlying cause of death, sex, age, country, and year from the World Health Organization Mortality Database,^9^ which compiles data from national vital registration systems. Population denominators were obtained from the Human Mortality Database.^10^

### Inclusion criteria

We included comparison countries that had a 2021 gross domestic product per capita greater than US $24,000 and were not formerly part of the Soviet Union or Eastern bloc, for consistency with prior work.^2^ Our analysis started in 1999, which is when the International Classification of Diseases, Tenth Revision (ICD-10) was adopted. We excluded countries that lacked mortality data for 2020 or failed to meet completeness and quality standards.^11^ This left 12 countries that satisfied our inclusion criteria (eTable 1 in the **Supplement**). Most had data for 1999 through 2020. The UK and Austria were missing data for 1999 to 2000 and 1999 to 2001, respectively, and were excluded in those years. Australia was missing data for 2005, and death counts were imputed using linear interpolation between adjacent years within age-by-sex-by-cause strata.

### Cause of death classification

All deaths were assigned to an ICD-10 code or to a residual (uncoded) category. Overall, 99.4% of US deaths (56.5 million) and 98.7% of peer country deaths (76.0 million) during the study period had a valid underlying cause of death recorded for analysis (eTable 1 in the **Supplement**). We aggregated ICD-10 codes into 17 mutually exclusive and collectively exhaustive cause-of-death categories, building on a classification by Elo et al.^12^ The categories ranked by frequency were circulatory diseases (e.g., heart disease, hypertension and stroke); other cancers (excluding lung cancer); mental and nervous system disorders (e.g., Alzheimer’s disease and related dementias, ADRD); respiratory diseases; lung cancer; diabetes, renal, and metabolic diseases; infectious and parasitic diseases; influenza and pneumonia; transportation accidents; drug poisonings; suicide; alcohol-related mortality; homicide; COVID-19; HIV/AIDS; symptoms, signs, and ill-defined conditions; and all other causes. eTable2 of the **Supplement** presents ICD-10 codes for these cause-of-death categories.

### Analysis

We calculated cause-specific mortality rates for each age group (0-4, 5-14, 15-24, 25-34, 35-44, 45-54, 55-64, 65-74, 75-84, 85+), country, and year by dividing cause-specific death counts by population denominators. We computed population-weighted average mortality rates in the 12 comparison countries for each age group, year, and cause-of-death category. We quantified the survival gap between the US and other wealthy nations using several metrics and assessed the contributions of different causes of death to this gap:

#### Excess US deaths

We quantified the number of US deaths that would have been expected if US mortality rates for a cause were equal to the average of peer nations. We calculated excess US deaths for each cause by subtracting the number of observed deaths from expected deaths.

#### Years of life lost (YLL)

We calculated YLL by weighting excess US deaths by the number of years the deceased would be expected to live if they had survived.^13^ These weights were equal to the average life expectancy of a resident of a peer country in the same age and sex group.

#### Ratio of observed to expected US deaths

To assess excess US mortality on a relative scale, we computed the ratio of cause-specific age-standardized mortality rates in the US compared to the weighted average of other wealthy nations. Mortality rates were standardized to the US age distribution in each year for consistency.

As there was no sampling, we do not present 95% confidence intervals. The study was determined not to be human subjects research by the Boston University Medical Campus Institutional Review Board (H-44819).

## RESULTS

From 1999 to 2020, 56.8 million deaths occurred in the US, and 77.0 million deaths occurred in 12 other wealthy nations (eTable 1 in the **Supplement**). The annual number of excess deaths in the US compared to peer countries increased from 365,599 in 1999 to 404,773 excess deaths in 2009, then 637,682 excess deaths in 2019, and 1,067,342 excess deaths in 2020 (**Table 1** and eTables 3-5 in the **Supplement**). Over the 21-year period, the US experienced 10,856,851 more deaths than would have occurred if the US had death rates equal to peer countries (eTable 6 in the **Supplement**).

**Table 1.** Excess deaths and excess YLL in the US compared to other wealthy nations by cause of death in 2019.

|  | Observed deaths | Counterfactual deaths | Ratio of observed to counterfactual | Excess deaths |  | Excess years of life lost |  |
| --- | --- | --- | --- | --- | --- | --- | --- |
|  |  |  |  | No. | % of total | No. | % of total |
| All-cause | 2 854 691 | 2 217 009 | 1.29 | 637 682 | 100 | 16 234 164 | 100 |
| Circulatory | 858 666 | 598 819 | 1.43 | 259 847 | 41 | 4 608 490 | 28 |
| Mental and nervous system disorders | 356 210 | 197 850 | 1.80 | 158 360 | 25 | 1 754 294 | 11 |
| Diabetes, renal, and metabolic | 175 627 | 82 621 | 2.13 | 93 006 | 15 | 1 883 240 | 12 |
| Respiratory | 221 236 | 144 209 | 1.53 | 77 027 | 12 | 1 329 350 | 8 |
| Drug poisoning | 70 622 | 10 558 | 6.69 | 60 064 | 9 | 2 418 474 | 15 |
| Transport accidents | 41 947 | 11 402 | 3.68 | 30 545 | 5 | 1 211 977 | 7 |
| Infectious and parasitic | 59 661 | 33 265 | 1.79 | 26 396 | 4 | 587 160 | 4 |
| Homicide | 18 998 | 1 444 | 13.16 | 17 554 | 3 | 860 579 | 5 |
| Alcohol-related | 39 034 | 26 163 | 1.49 | 12 871 | 2 | 448 249 | 3 |
| Lung cancers | 139 680 | 129 687 | 1.08 | 9 993 | 2 | 150 329 | 1 |
| Suicide | 42 684 | 32 982 | 1.29 | 9 702 | 2 | 442 071 | 3 |
| HIV/AIDS | 5 042 | 770 | 6.55 | 4 272 | 1 | 133 090 | 1 |
| Influenza and pneumonia | 49 780 | 88 918 | 0.56 | –39 138 | –6 | –267 747 | –2 |
| Other cancers | 475 545 | 522 957 | 0.91 | –47 412 | –7 | –384 696 | –2 |
| Symptoms, signs, and ill-defined conditions | 47 671 | 124 369 | 0.38 | –76 698 | –12 | –719 440 | –4 |
| All other causes | 252 288 | 210 996 | 1.20 | 41 292 | 6 | 1 778 743 | 11 |

**Abbreviations:** YLL = Years of Life Lost**Notes:** The counterfactual refers to the expected number of deaths the US would have experienced if it had death rates equal to the average of 12 other wealthy nations, which are described in eTable 1. Results for 2019 are shown because it was the most recent year prior to the COVID-19 pandemic.

### US Excess Mortality by Cause of Death in 2019

**Table 1** decomposes excess US deaths by cause of death in 2019, the most recent year prior to the COVID-19 pandemic. Of the 637,682 excess US deaths in 2019, 259,847 (41% of total) were caused by circulatory diseases and 158,360 (25%) by mental and nervous system disorders. Diabetes, renal, and metabolic conditions contributed 93,006 excess deaths (15%). An additional 82,637 excess deaths (13%) were due to drug poisonings (60,064), alcohol (12,871), and suicide (9,702). Respiratory conditions contributed 77,027 (12%) and transportation accidents contributed 30,545 (5%) excess deaths. Compared to peer countries, the US had fewer deaths from other cancers and from symptoms, signs, and ill-defined conditions.

Excess US deaths in 2019 were responsible for 16,234,164 YLL, a measure that places greater weight on deaths earlier in life (**Figure 1**). Circulatory disease contributed less to excess YLL than to excess deaths (28% of excess YLL vs. 41% of excess deaths) as did mental and nervous system disorders (11% vs. 25%). In contrast, deaths due to drug poisonings, alcohol, and suicide contributed more to YLL than to excess deaths (20% vs. 13%) and at 3,308,794 were the second leading cause of YLL.

**Figure 1.**
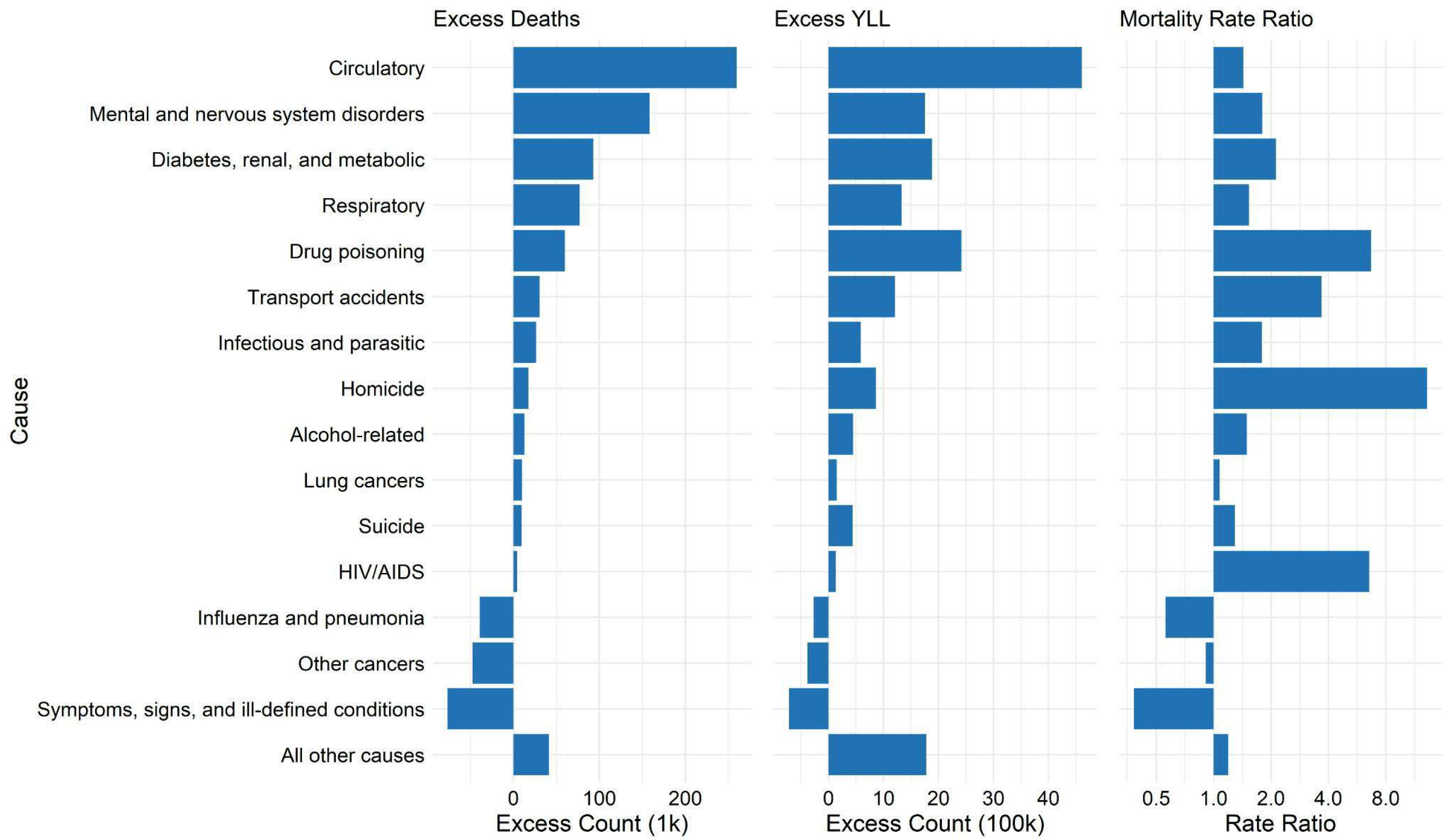
Excess deaths and excess YLL in the US compared to peer countries by cause of death in 2019 **Abbreviations:** YLL = Years of Life Lost **Notes:** The mortality rate ratio refers to the ratio of the age-standardized death rate for a cause of death in the US compared to other wealthy nations. It is a relative measure of the extent to which US mortality rates exceed rates in other wealthy nations. The 12 other wealthy nations are described in eTable 1. Results for 2019 are shown because it is the most recent year prior to the COVID-19 pandemic. This figure is a visualization of data from Table 1.

Causes contributing the most excess deaths differed from those with the largest relative rates. **Table 1** and **Figure 1** present ratios of observed to expected US deaths for each cause, which can be interpreted as age standardized rate ratios. Death rates for homicide and HIV/AIDS were 13.2 times and 6.6 times higher in the US compared to peer countries. Despite these large relative differences, these causes were responsible for 3% and 0.7% of excess deaths, respectively. On the other hand, circulatory death rates were 1.4 times higher in the US but were the leading cause of excess deaths in 2019. Diabetes, renal, and metabolic mortality was 2.1 times higher in the US than peer countries, transportation deaths 3.7 times higher, and drug poisonings 6.7 times higher.

### Changes in US Excess Deaths by Cause of Death from 1999 to 2019

**Figure 2** shows annual excess deaths and YLL in the US by cause of death from 1999 to 2020, highlighting comparisons between causes (top panel) and their relative contribution to the total excess (bottom panel). eFigure 1 and eFigure 2 in the **Supplement** show observed, expected, and excess deaths and YLL for the full period. eTables 3-5 in the **Supplement** present excess deaths, excess YLL, and rate ratios by cause of death for 1999, 2009, and 2020, complementing the 2019 data in **Table 1**.

**Figure 2.**
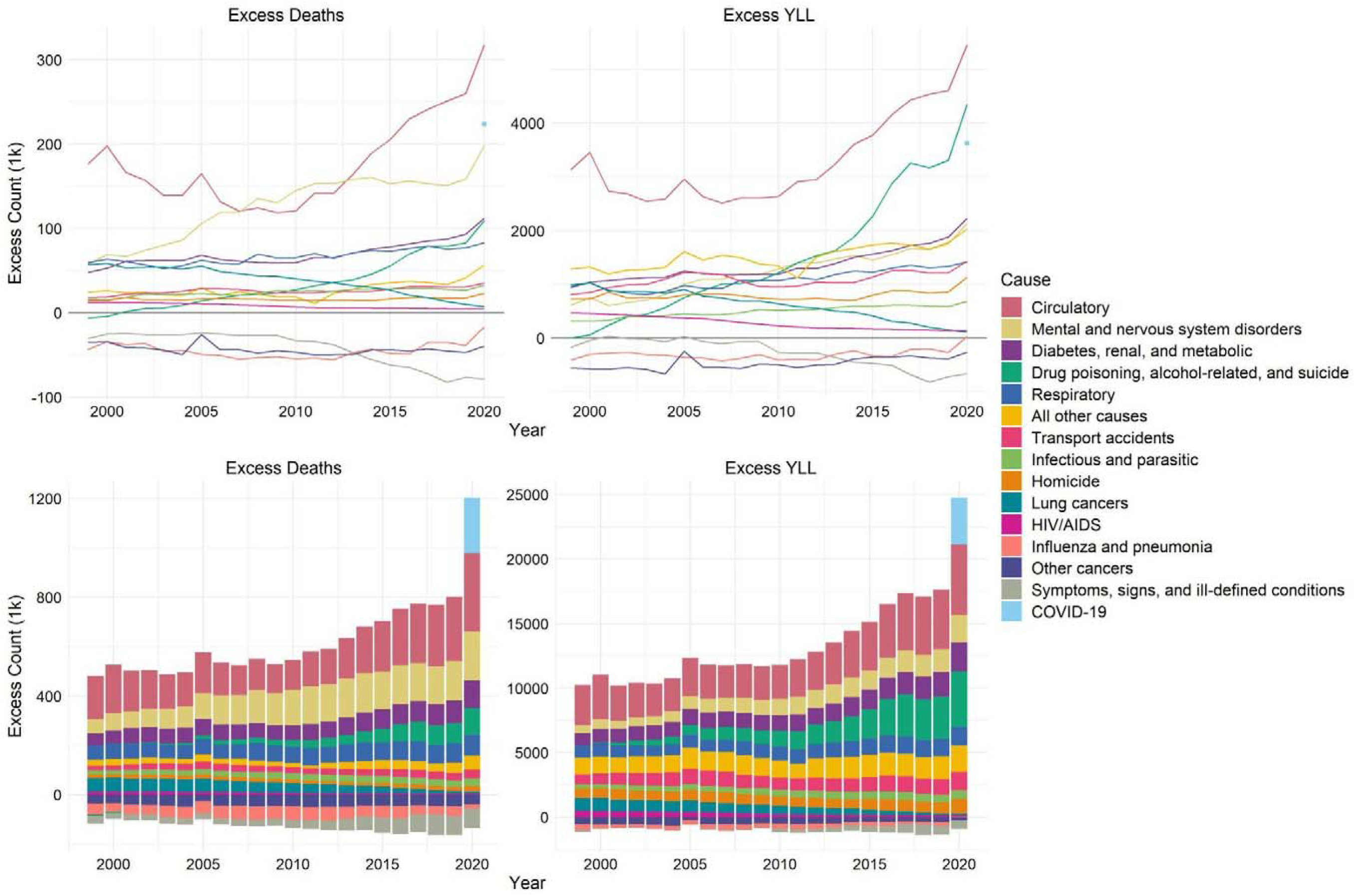
Annual excess deaths and excess YLL in the US compared to other wealthy nations by cause of death from 1999 to 2020 **Abbreviations:** YLL = Years of Life Lost **Notes:** The top panels plot the annual number of excess deaths and YLLs for each cause of death, whereas the bottom panels are stacked bar charts decomposing the total annual number of excess deaths and YLLs by cause of death to demonstrate the contribution of each cause of death to the total excess.

Circulatory diseases and mental and nervous system conditions were the leading causes of excess deaths in the US compared to other wealthy nations each year (**Figure 2**). Annual excess deaths from circulatory disease fell from 176,499 in 1999 to 130,127 in 2009, before rising to 259,847 in 2019. From 2009 to 2019, circulatory mortality was the largest contributor to the growth in excess deaths and YLL (**Figure 3**). eFigure 1 in the **Supplement** shows that US circulatory deaths declined along with peer countries until around 2010 when they stagnated and then increased, diverging from peer countries. Excess deaths due to diabetes, renal, and metabolic disease, which have some overlapping risk factors with circulatory disease, increased slowly from 1999-2009 and more rapidly from 2009-2019 when these causes were responsible for the second-largest increase in excess deaths.

**Figure 3.**
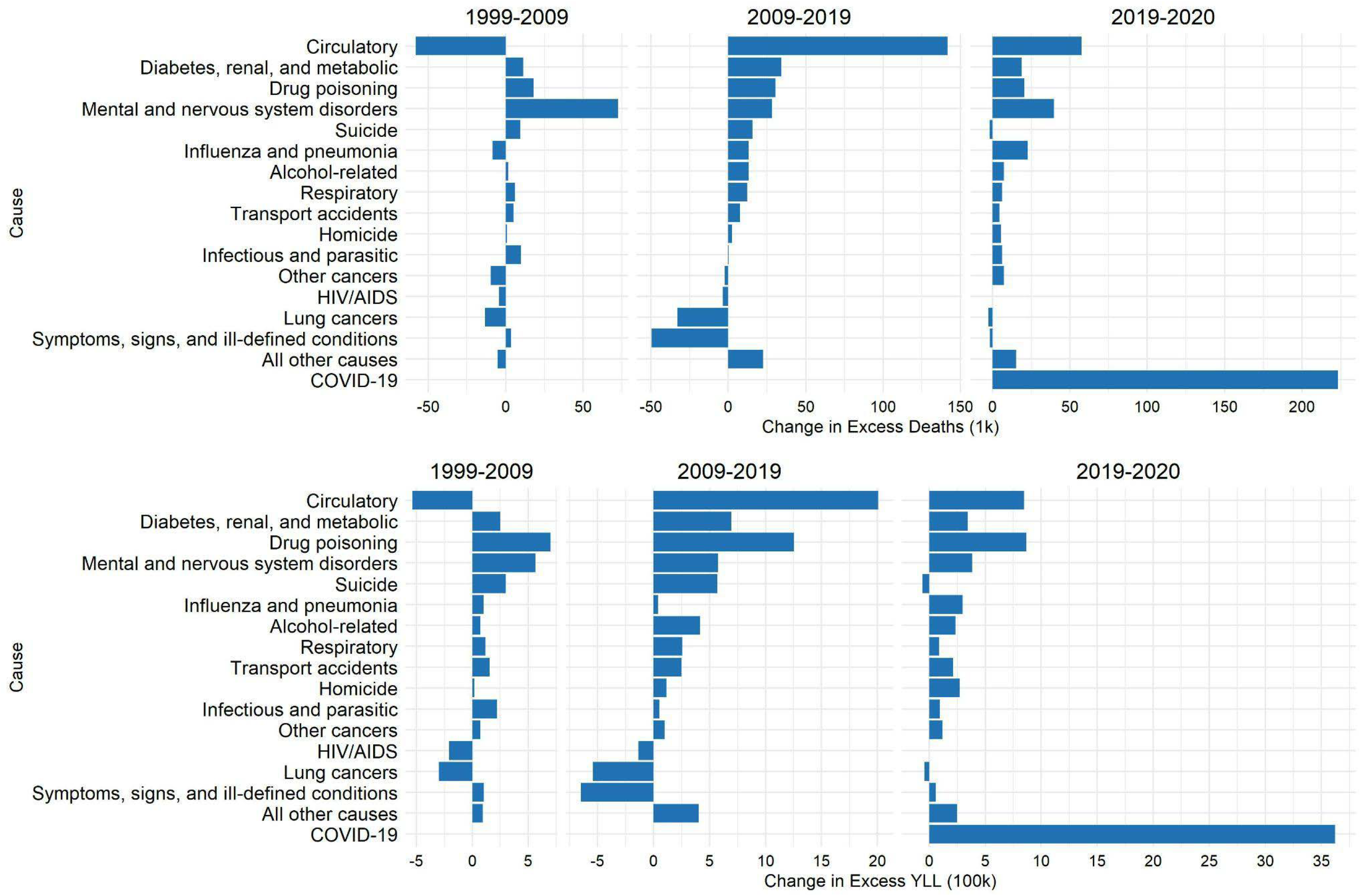
Changes in excess deaths and excess YLL in the US compared to other wealthy nations over 3 time intervals (2009 compared to 1999, 2019 compared to 2009, and 2020 compared to 2019) by cause of death **Abbreviations:** YLL = Years of Life Lost

Excess deaths from mental and nervous system disorders increased rapidly from 57,511 in 1999 to 130,127 in 2009, and then plateaued, reaching 158,360 in 2019 (**Figure 2**), a 2.8-fold increase over the study period. Mental and nervous system disorders had the largest increase in excess deaths of any cause during 1999-2009 (**Figure 3**). Their contribution to the growth in YLL was less pronounced because such deaths often occurred at older ages. Drug poisonings, often at younger ages, were responsible for the largest increase in YLL in the first decade.

Excess deaths from drug poisonings, alcohol, and suicide in the US compared to other wealthy nations increased steadily over the period. In 1999, there were more of these deaths in other wealthy nations than in the US because of lower US suicide rates (eTable 3 in the **Supplement**). However, excess deaths soon increased for all three causes, with the largest increase in deaths from drug poisonings. From 1999 to 2019, annual excess deaths due to drug poisonings, alcohol, and suicide increased by 88,574 deaths (48,643 deaths from drug poisoning; 14,747 deaths from alcohol-induced causes; 25,183 deaths from suicide). These causes represented 13% of US excess deaths in 2019 but 33% of the increase in excess deaths and 46% of the increase in excess YLL between 1999 and 2019 (**Table 1**, eTable 3 in the **Supplement**).

Excess deaths due to lung cancer decreased from 56,440 in 1999, when it was the second-leading cause, to 9,993 excess deaths in 2019.

### Changes in US Excess Deaths by Cause of Death from 2019 to 2020

**Figure 3** quantifies changes in excess deaths between 2019 and 2020, the first year of the COVID-19 pandemic. In 2020, the mortality disadvantage for the US compared to other wealthy nations increased by 429,659 deaths, a markedly larger increase than between 2018 and 2019 when excess deaths increased by 31,055 deaths. Of the additional excess deaths in the US in 2020, 52% (223,398 deaths) had COVID-19 listed as the underlying cause of death (eTable 7 in the **Supplement**). Though COVID-19 led to a large increase in mortality across countries, COVID-19 only accounted for 21% of excess deaths (and 15% of excess YLL) in the US relative to peer countries in 2020. Circulatory diseases contributed most to excess deaths in 2020, and circulatory diseases followed by drug poisonings, alcohol-related deaths, and suicide accounted for the most YLL in 2020 (**Figure 2**, eTable 5 in the **Supplement**).

### Age Patterns in US Excess Deaths by Cause of Death

**Figure 4** examines age-specific patterns in the causes of excess deaths and YLL in 2019. Transportation accidents, homicides, and drug poisonings, alcohol-related mortality, and suicide explained most excess deaths among persons at ages 0-24 years. Drug poisonings, alcohol-related deaths, and suicide also caused a large number of excess US deaths at ages 25-34, 35-44, 45-54, and 55-64, and were the leading cause of excess deaths at ages 25-44. In 2019, more than half of excess deaths in the US compared to peer countries for adults at ages 25-44 were related to these causes (eTable 8 in the **Supplement**).

**Figure 4.**
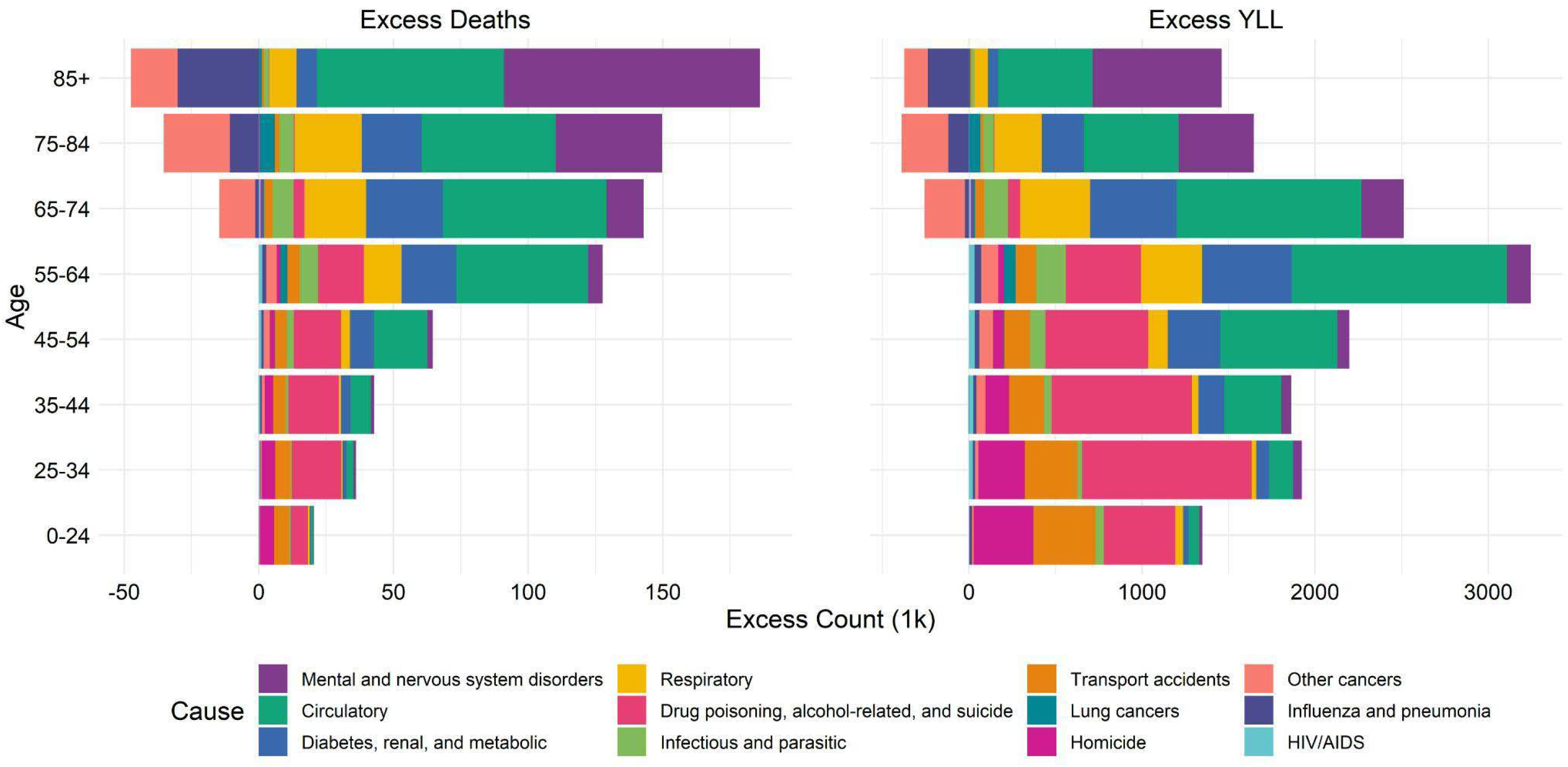
Age differences in excess deaths and excess YLL in the US compared to peer countries by cause of death in 2019 **Abbreviations:** YLL = Years of Life Lost | | HIV/AIDS = Human Immunodeficiency Virus / Acquired Immunodeficiency Syndrome

Circulatory diseases were the leading cause of death for all age groups over 45 years. Excess deaths due to diabetes, renal, and metabolic conditions were also common at these ages. Taken together, excess deaths due to circulatory and metabolic conditions were equal to over half of all excess deaths for persons ages 55-64, 65-74, and 75-84. Respiratory conditions had an increasing role in excess mortality starting at age 55. Mental and nervous system disorders emerged as a common cause of excess deaths starting at ages 65 years and older and were the leading cause of excess deaths for persons ages 85 and older.

eFigure 3 and eFigure 4 in the **Supplement** show how cause-specific excess deaths and YLL changed from 1999 to 2020 in different age groups. The rise in US excess deaths for persons aged 0 to 44 years was due *entirely* to increases in drug poisonings, alcohol-related mortality and suicide, relative to peer countries. This increase became steeper in 2014, stabilized from 2017 to 2019, and increased again in 2020. For persons aged 45-64 years, the rise in excess deaths was driven by a steep rise in drug poisonings, alcohol, and suicide; a steady rise in circulatory mortality; a continuous rise in deaths from diabetes, renal, and metabolic conditions; and, in 2020, by COVID-19. For persons ages 65 and older, the rise in excess deaths was driven in the first decade (1999-2009) by an increase in mental and nervous system conditions, in the second decade (2010-2020) by an increase in circulatory mortality, and in 2020 by COVID-19. Excess circulatory deaths exhibited markedly different trends for persons aged 45-64 years and ages 65 and older, suggesting cohort-specific patterns in circulatory mortality.

## DISCUSSION

The US had substantially higher mortality rates than other wealthy nations from 1999 to 2020,^3,14–16^ with hundreds of thousands of “missing Americans” each year.^2,17^ The US mortality disadvantage predates the COVID-19 pandemic but increased during it. Circulatory conditions (e.g., heart disease and stroke) were responsible for the largest share of excess US deaths and YLL in nearly every year from 1999 to 2020. Since around 2010, the decline in circulatory mortality stalled in the US but continued in peer nations, leading to an increase in excess US deaths. Mehta et al. showed that if the US decline in cardiovascular mortality had continued, US life expectancy would have kept pace with peers during the 2010s.^7^ For ages 45 to 64, we found that US circulatory mortality started diverging from other wealthy nations as early as 2001, whereas for ages 65 and older the US circulatory mortality disadvantage first narrowed before widening after 2010.

The largest contributor to the growth in excess US deaths between 1999 and 2019 was the increase in drug poisonings, alcohol-related mortality, and suicide, often conceptualized as “deaths of despair”.^18–22^ The US had death rates similar to peers for these causes in 1999. However, by 2019 the US experienced nearly 83,000 excess deaths due to drugs, alcohol, and suicides each year. With respect to drug poisonings, the US was a significant outlier,^5^ with drug poisonings in the U.S. occurring nearly 7 times more frequently than other wealthy nations.

Some major causes of excess US deaths also had large mortality rate ratios relative to peer countries, suggesting substantial scope to reduce US mortality with existing technologies and interventions. Diabetes, renal, and metabolic conditions were a substantial and growing contributor, consistent with prior evidence that the US faces elevated morbidity and mortality associated with cardiometabolic risk factors, including obesity, hyperglycemia, and poor kidney function.^23–25^ Mental and nervous system disorders (e.g., ADRD) were also a major contributor, particularly at ages 85 and older. Transportation accidents constituted a large and growing share of excess US deaths. Finally, death rates due to homicide and HIV/AIDS were both higher in the US than in peer countries and continued to diverge over time but were smaller contributors to the absolute number of excess US deaths.

Death certificates identify proximate causes of death such as diseases or external factors. Our findings therefore must be assessed alongside literature on upstream social, economic, environmental, and political factors that give rise to differential mortality rates.^26–31^ Several studies have linked mortality outcomes in the US with social and economic changes, including trade liberalization, the loss of manufacturing jobs, automation, and worsened opportunities for lower-educated workers.^31–34^ US regions with less protective safety net and health coverage policies have higher and worsening mortality than other regions,^28,35^ as do those where majorities voted for Donald Trump in 2016.^17,36^ Other studies have explored supply-side causes of mortality trends,^37^ including the over-prescription of opioids starting in the late 1990s,^38^ the proliferation of fentanyl since 2013;^5^ and the rise of obesity and consequent metabolic disease associated with changes in the food environment.^24^

Our findings suggest a need for research to address why US trends in circulatory disease deaths have diverged from trends in peer countries; why mental and nervous system mortality has increased faster in the US than other wealthy nations; why US drug poisonings have increased at all ages and especially among young people; and to what extent changes in firearm, transit, and nutrition policies can reduce US excess mortality due to homicide and suicide, transportation, and cardiometabolic diseases.

Our study had several limitations. First, diagnostic and coding practices may vary between countries and over time. Second, peer countries had higher death rates reported to unspecified causes, particularly later in the study period. This may reflect reporting lags which, when resolved, will increase deaths reported to other categories. Third, we followed prior practice^2^ in choosing our counterfactual to estimate excess US deaths. Comparisons with more nations, which may become feasible as more countries report cause-disaggregated data may yield different estimates. Finally, death certificates do not assign causality to important determinants of mortality such as social disadvantage, racism and other structural determinants of health, and policy regimes, factors which are likely to be critical targets for intervention.^39^

From 1999 to 2020, 10.8 million Americans were missing due to US mortality rates exceeding mortality rates in other wealthy nations. This “population autopsy” examined the causes of death responsible for these missing Americans. Although other wealthy nations have access to similar advanced medical technologies as the US, they have enjoyed far better health outcomes, likely due in part to different health and social policy choices. This analysis suggests avenues for future research and policy development to address the growing US mortality disadvantage for specific causes of death.

## Data Availability

All data used in this analysis are publicly available. Study data and analytic code used to generate all estimates are posted in a public data repository.

https://osf.io/85sk2/

## AUTHOR CONTRIBUTIONS

JB, DS, SW, and ACS conceived the analysis and designed the study. RR extracted the data and conducted the analysis under the supervision of JB and ACS. JB wrote the first draft of the manuscript. All authors critically revised the manuscript.

## NON-AUTHOR CONTRIBUTIONS

The authors would like to thank Shaylan Hill for her assistance with data cleaning and analysis.

## ACCESS TO DATA AND ANALYSIS

JB had full access to all the data in the study and takes responsibility for the integrity of the data and the accuracy of the data analysis.

## DATA SHARING STATEMENT

All data used in this analysis are publicly available. Study data and analytic code used to generate all estimates are posted in a public data repository (https://osf.io/85sk2/).

## MEETING PRESENTATION

This work was presented at the Interdisciplinary Association for Population Health Science annual meeting in Baltimore, MD, in October 2023 and at the Population Association of America annual meeting in Columbus, Ohio, in April 2024.

