## Supplement for "Causes of excess deaths in the US relative to other wealthy nations, 1999-2020: a population autopsy"

### Contents:

**eTable 1.** Total deaths and person-time in the US and 12 other wealthy nations by cause of death from 1999 to 2020

**eTable 2.** ICD-10 code typology for cause of death categories used in this analysis

**eTable 3.** Excess deaths and excess YLL in the US compared to other wealthy nations by cause of death in 1999

**eTable 4.** Excess deaths and excess YLL in the US compared to other wealthy nations by cause of death in 2009

**eTable 5.** Excess deaths and excess YLL in the US compared to other wealthy nations by cause of death in 2020

**eTable 6.** Excess deaths and excess YLL in the US compared to other wealthy nations by cause of death from 1999 to 2020

**eTable 7.** Change in excess deaths and excess YLL in the US compared to other wealthy nations by cause of death from 1999 to 2020

##### eTable 8. Excess deaths and excess YLL in the US compared to other wealthy nations by age and cause of death in 2019

##### eFigure 1. Annual observed deaths in the US, counterfactual deaths in other wealthy nations, and excess deaths in the US compared to other wealthy nations for each cause of death from 1999 to 2020

##### eFigure 2. Annual observed YLL in the US, counterfactual YLL in other wealthy nations, and excess YLL in the US compared to other wealthy nations for each cause of death from 1999 to 2020

**eFigure 3.** Annual excess deaths by age-group in the US compared to other wealthy nations by cause of death from 1999 to 2020

**eFigure 4.** Annual excess deaths and excess YLL by age-group in the US compared to other wealthy nations by cause of death from 1999 to 2020

**Supplementary Data** & **Replication Code**

##### eTable 1. Total deaths and person-time in the US and 12 other wealthy nations by cause of death from 1999 to 2020

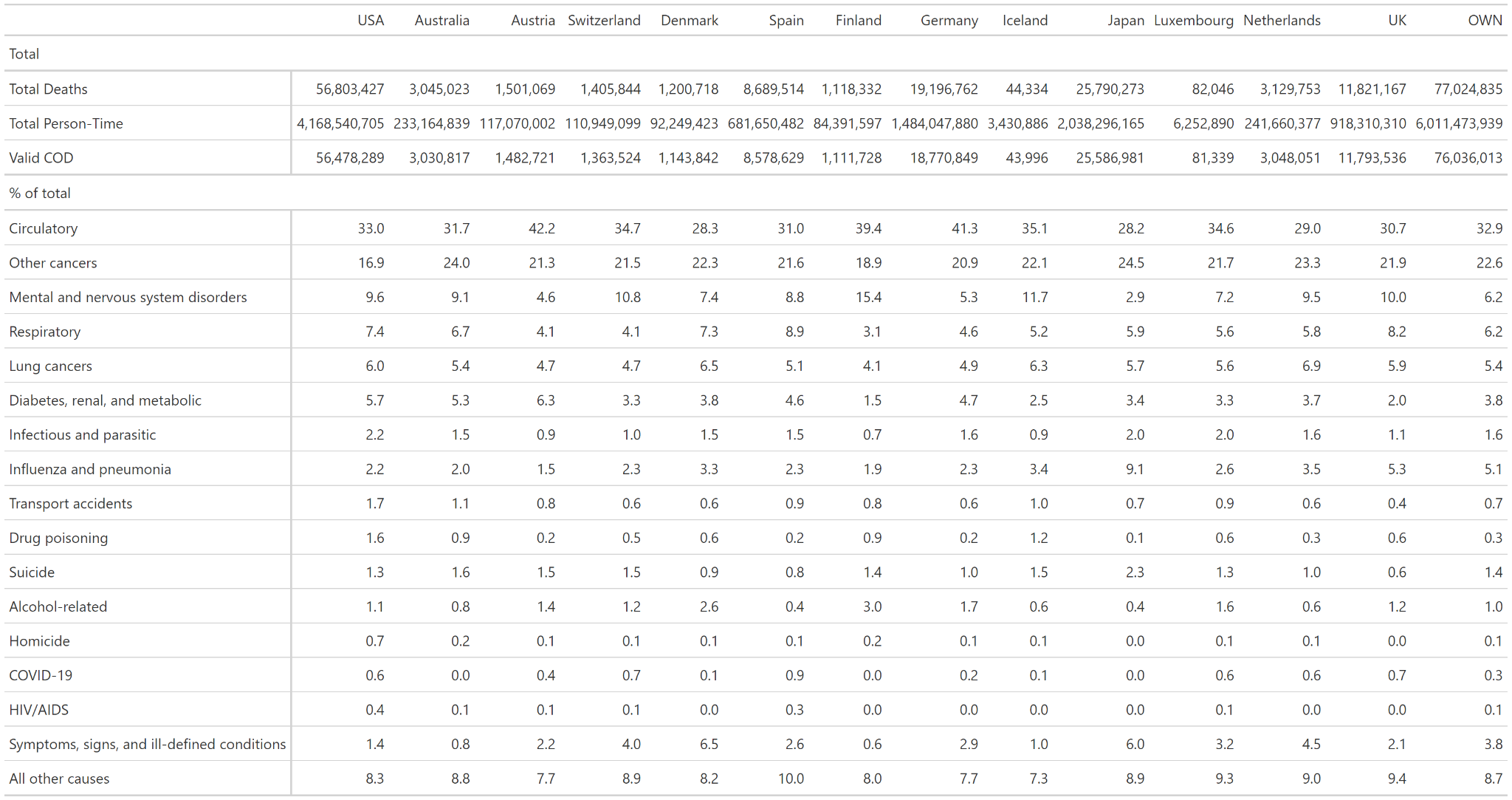

**Abbreviations:** OWN = Other wealthy nations

**Notes:** OWN is the aggregate of all comparison countries

##### eTable 2. ICD-10 code typology for cause of death categories used in this analysis

| **Categories** | **ICD-10 Codes** |
| --- | --- |
| Circulatory diseases | I00–I99 (excluding I42.6) |
| Other cancers (excluding lung cancers) | C00–D48 (excluding C33, C34) |
| Mental and nervous system disorders^a^ | F01–F99 (excluding F10), G00–G98 (excluding G31.2, G62.1, G72.1) |
| Respiratory diseases (excluding influenza and pneumonia) | J00–J98 (excluding J09–J18) |
| Lung cancers | C33–C34 |
| Diabetes, renal, and metabolic diseases | E10–E14, N17–N19, E7–E88, E65–E68 |
| Infectious and parasitic diseases | A00–B99 (excluding B20–B24) |
| Influenza and pneumonia | J09–J18 |
| Transport accidents | V01–V99, Y85 |
| Drug poisoning^b^ | X40–X44, X60–X64, X85, Y10–Y14 |
| Suicide | X60–X84, Y87.0 (excluding X60–X65) |
| Alcohol-related causes | E24.4, F10, G31.2, G62.1 G72.1, I42.6, K29.2, K70, K85.2, K86.0, R78.0, X45, X65, Y15 |
| Homicide^c^ | X86–Y09, Y87.1 |
| COVID-19 | U07.1 |
| HIV/AIDS | B20–B24 |
| Symptoms, signs, and ill-defined conditions | R00–R99 (excluding R78.0) |
| All other causes | D50–D89, E00–E90 (excluding E10–E14, E65–E68, E70–E88), G99, H00–H93, J99, K00–K92, L00–L98, M00–M99, N00–N99 (excluding N17–N19), O00–O99, P00–P96, Q00–Q99, U00–U99, V01–Y89 (excluding X40–X44, X45, X60–Y15, Y87.0–Y87.1), Y90–Y98 |

^a^Includes Alzheimer’s disease and related dementias (ADRD).

^b^Except suicide by drugs, which is included in drug overdose.

^c^Except assault by drugs, medicaments, and biological substances, which is included in drug overdose.

**Notes:** This list is ordered by the number of deaths that occurred in the US from 1999 to 2020, besides symptoms, signs and ill-defined conditions, and all other causes. To modify the list from Elo et al., we added the following cause-of-death categories: transport accidents, COVID-19, and infectious and parasitic disease. We expanded the breast, prostate, colorectal, and cervical cancer category to include all other cancers besides lung cancers, and the diabetes category to include renal and metabolic diseases.

##### eTable 3. Excess deaths and excess YLL in the US compared to other wealthy nations by cause of death in 1999

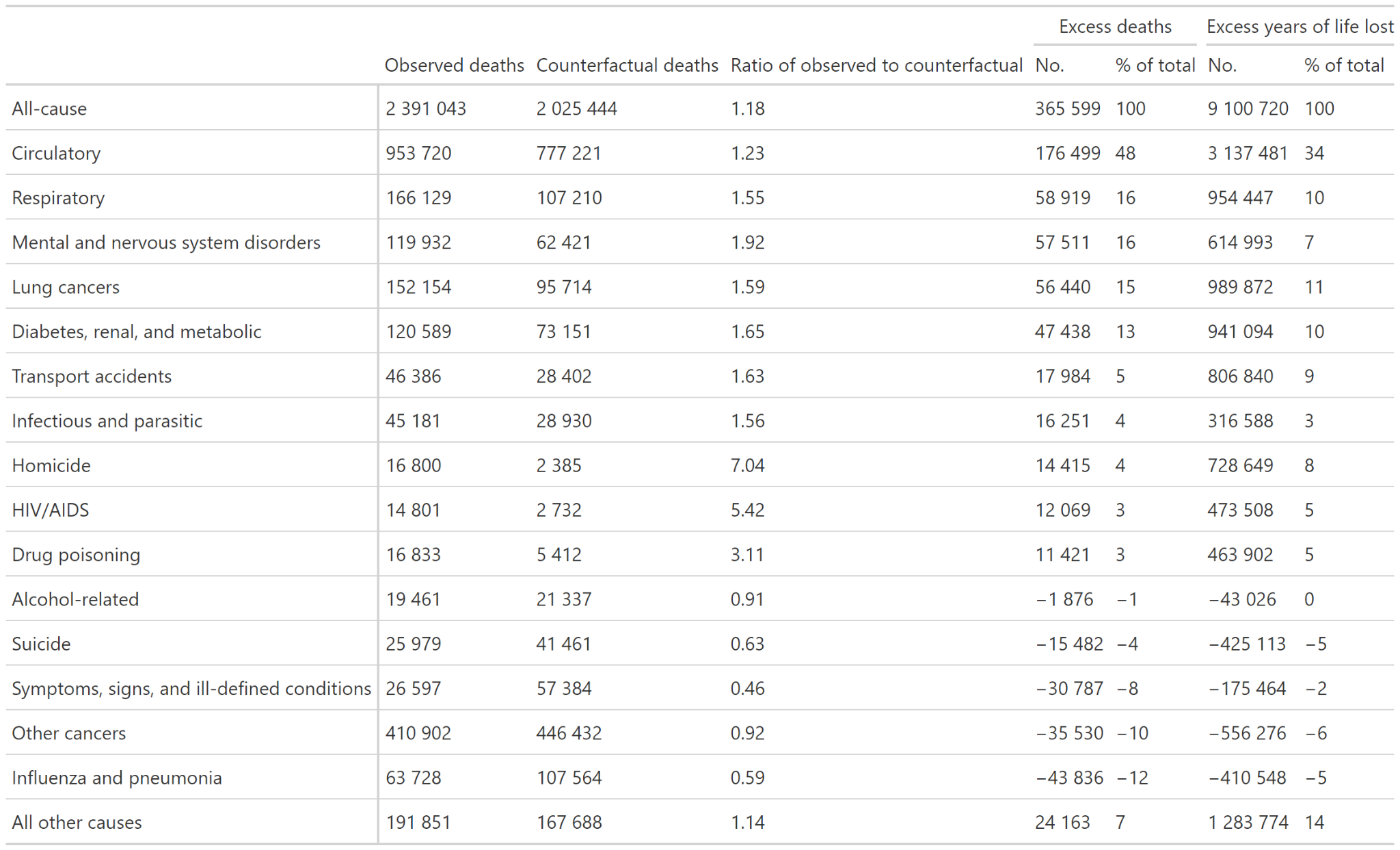

**Abbreviations:** YLL = Years of Life Lost

**Notes:** The counterfactual refers to the expected number of deaths the US would have experienced if it had death rates equal to the average of 12 other wealthy nations, which are described in eTable 1.

##### eTable 4. Excess deaths and excess YLL in the US compared to other wealthy nations by cause of death in 2009

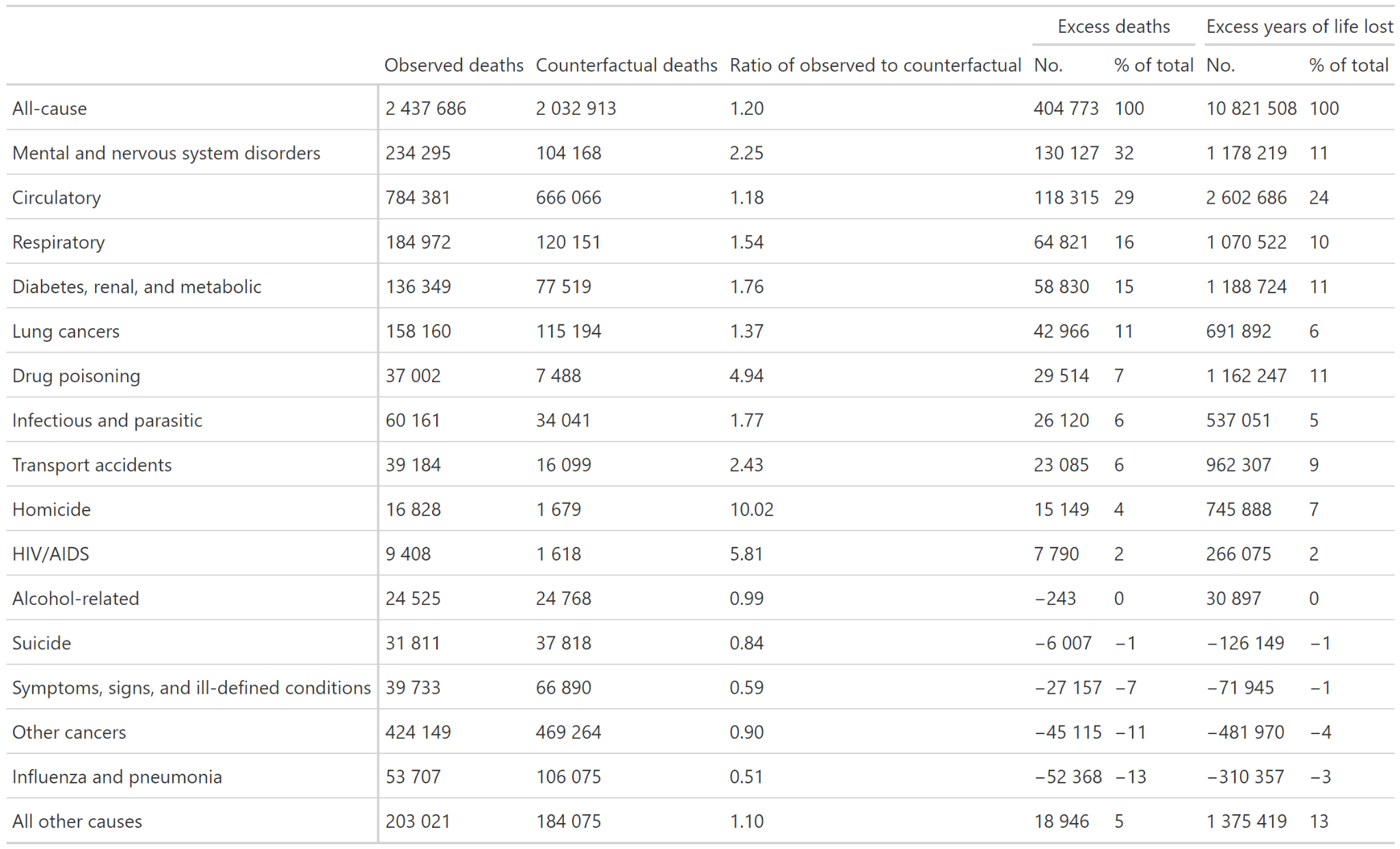

**Abbreviations:** YLL = Years of Life Lost

**Notes:** The counterfactual refers to the expected number of deaths the US would have experienced if it had death rates equal to the average of 12 other wealthy nations, which are described in eTable 1.

##### eTable 5. Excess deaths and excess YLL in the US compared to other wealthy nations by cause of death in 2020

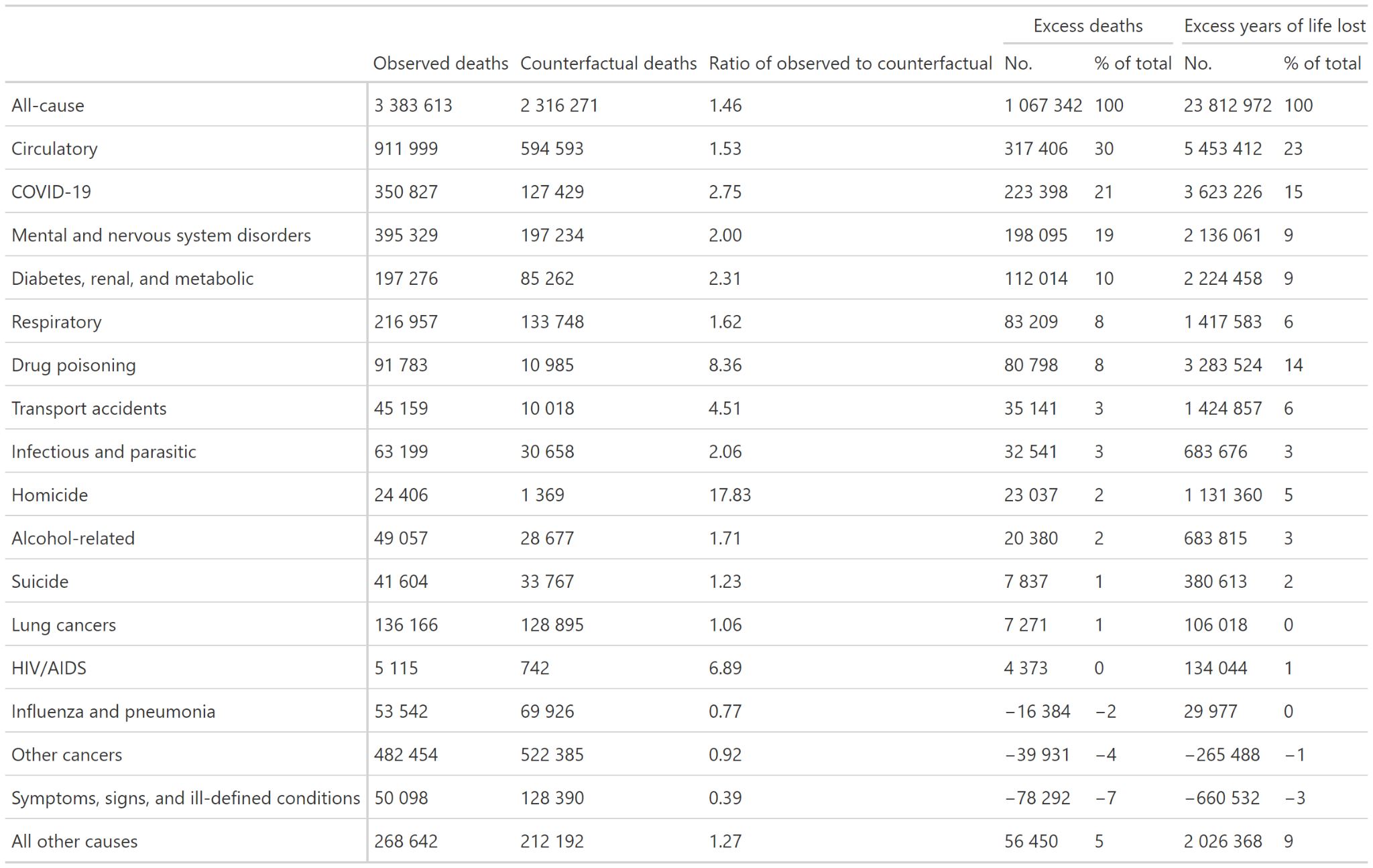

**Abbreviations:** YLL = Years of Life Lost

**Notes:** Notes: The counterfactual refers to the expected number of deaths the US would have experienced if it had death rates equal to the average of 12 other wealthy nations, which are described in eTable 1. Results for 2020 reflect the only year that sufficient data were available during the COVID-19 pandemic.

##### eTable 6. Excess deaths and excess YLL in the US compared to other wealthy nations by cause of death from 1999 to 2020

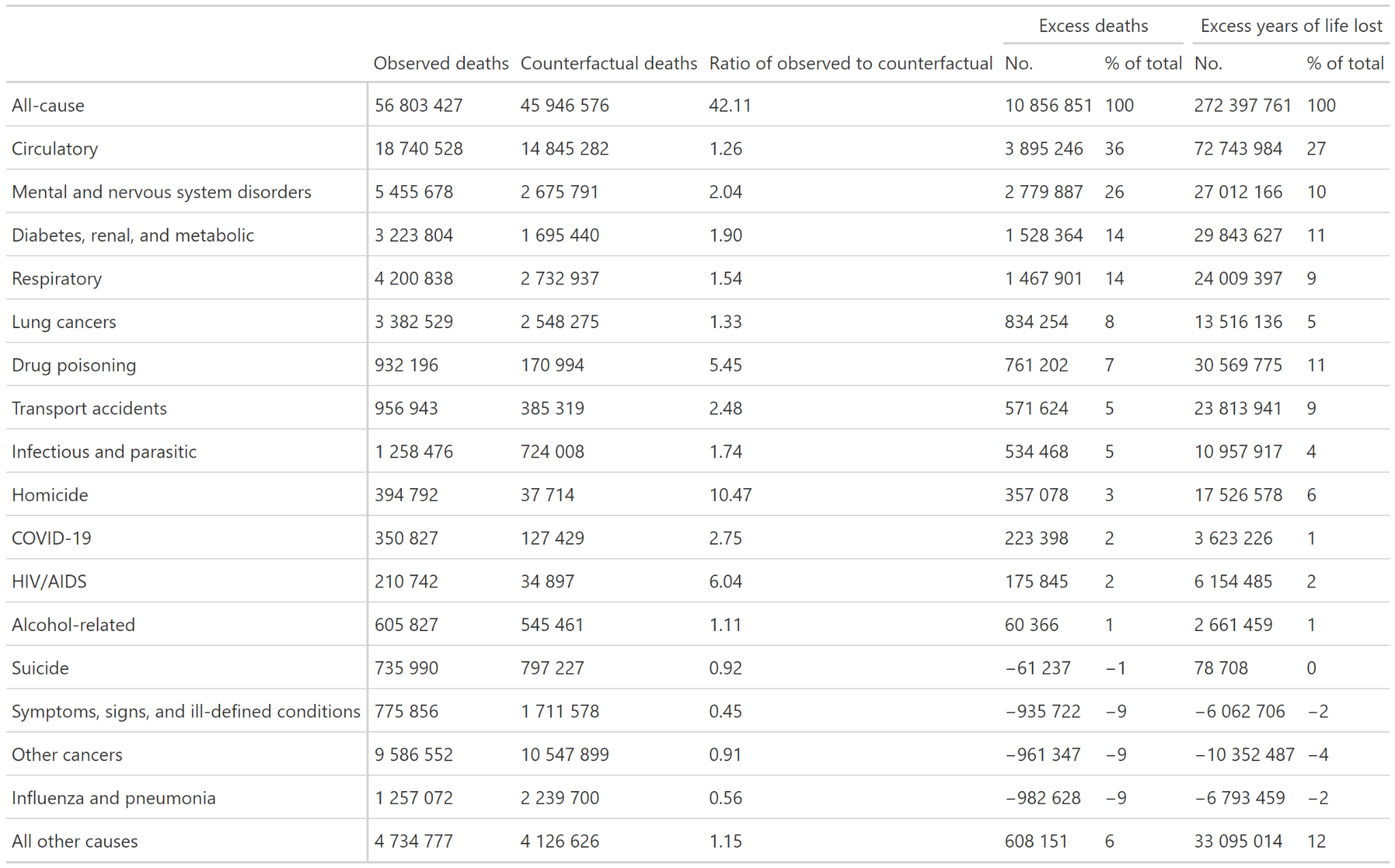

**Abbreviations:** YLL = Years of Life Lost

**Notes:** Notes: The counterfactual refers to the expected number of deaths the US would have experienced if it had death rates equal to the average of 12 other wealthy nations, which are described in eTable 1. Results for 2020 reflect the only year that sufficient data were available during the COVID-19 pandemic.

##### eTable 7. Changes in excess deaths and YLL in the US compared to other wealthy nations by cause of death from 1999 to 2019 and 2019 to 2020

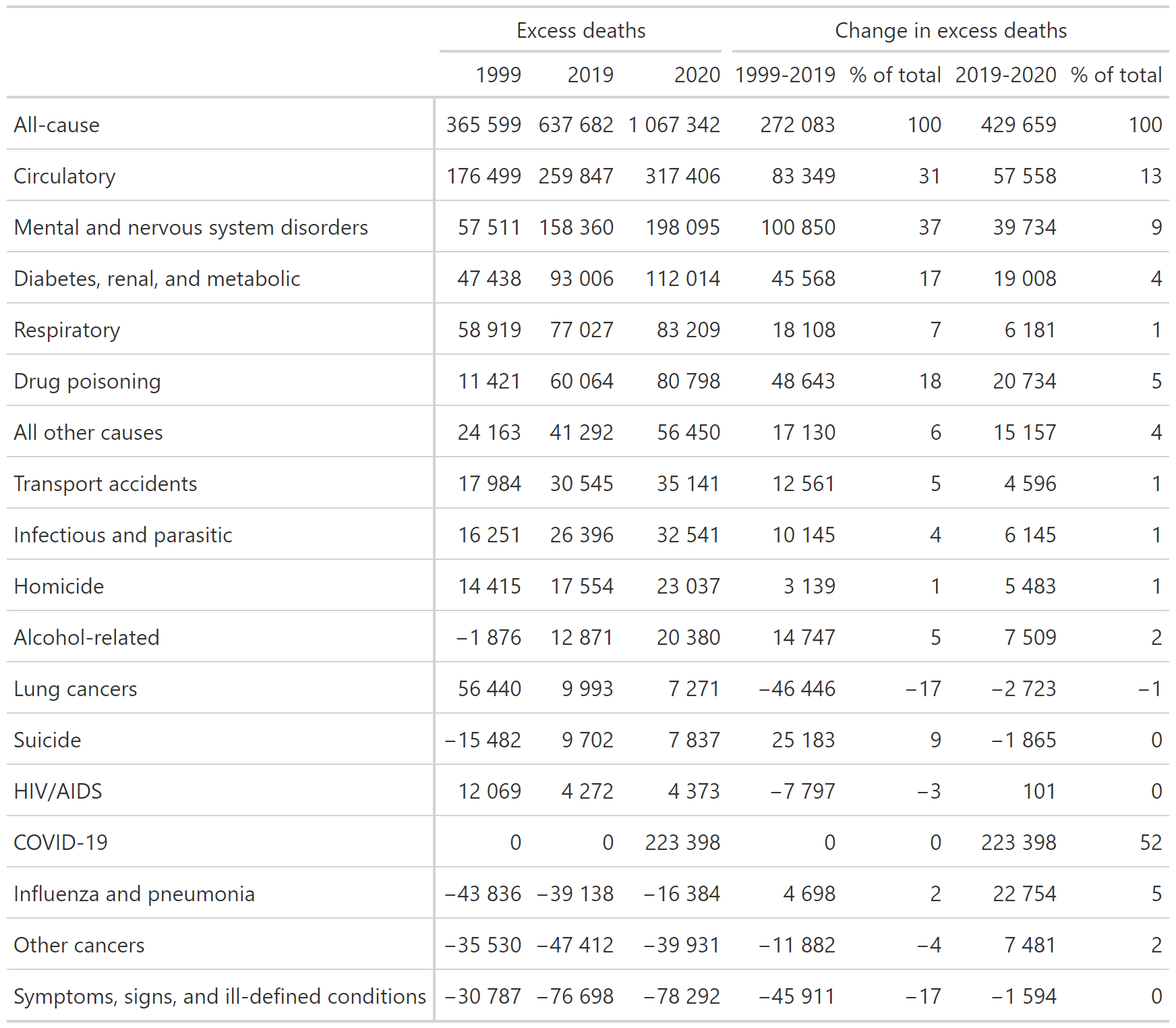

##### eTable 8. Excess deaths and excess YLL in the US compared to other wealthy nations by age and cause of death in 2019

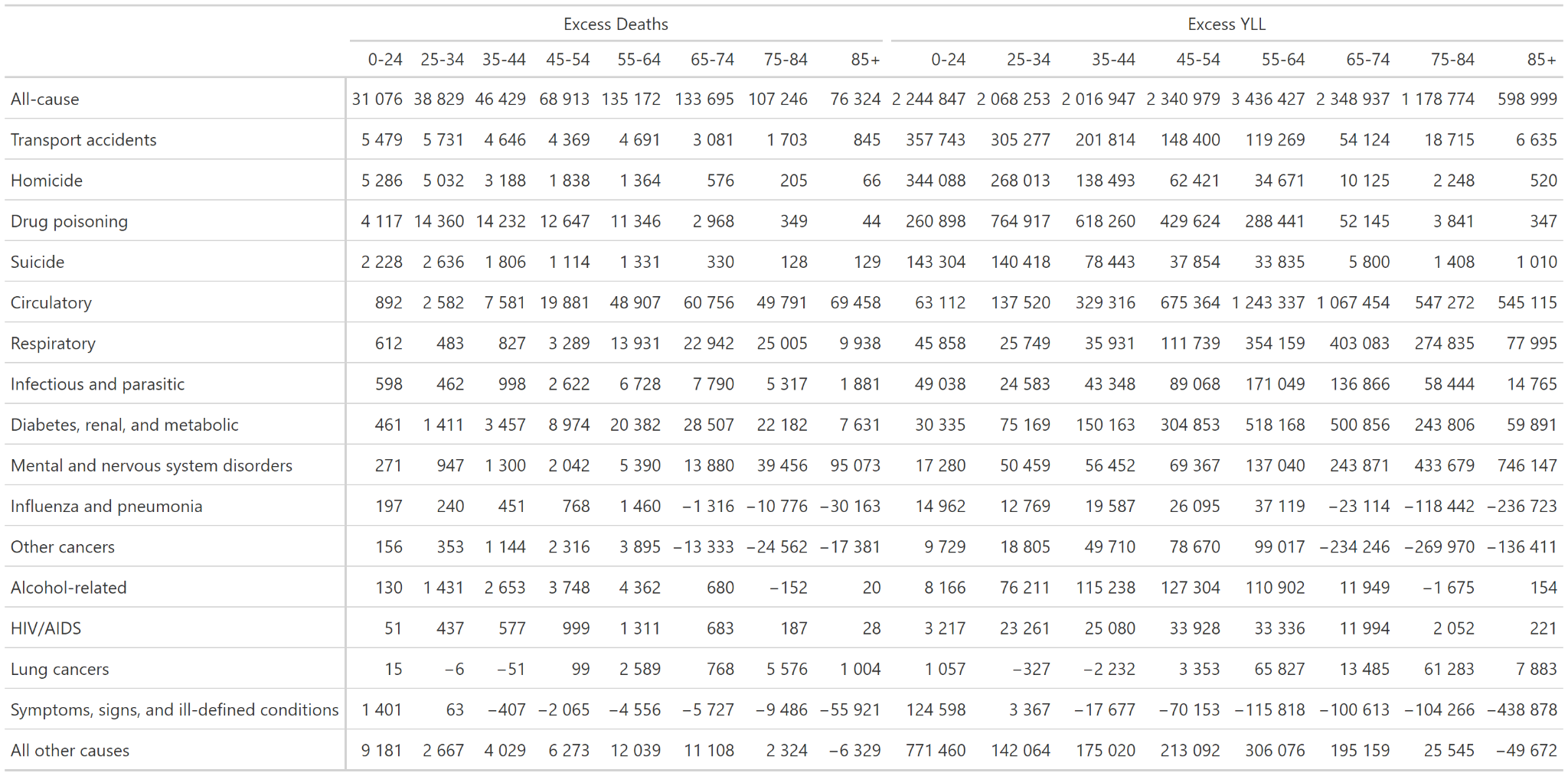

**eFigure 1.** Annual observed deaths in the US, counterfactual deaths in other wealthy nations, and excess deaths in the US compared to other wealthy nations for each cause of death from 1999 to 2020

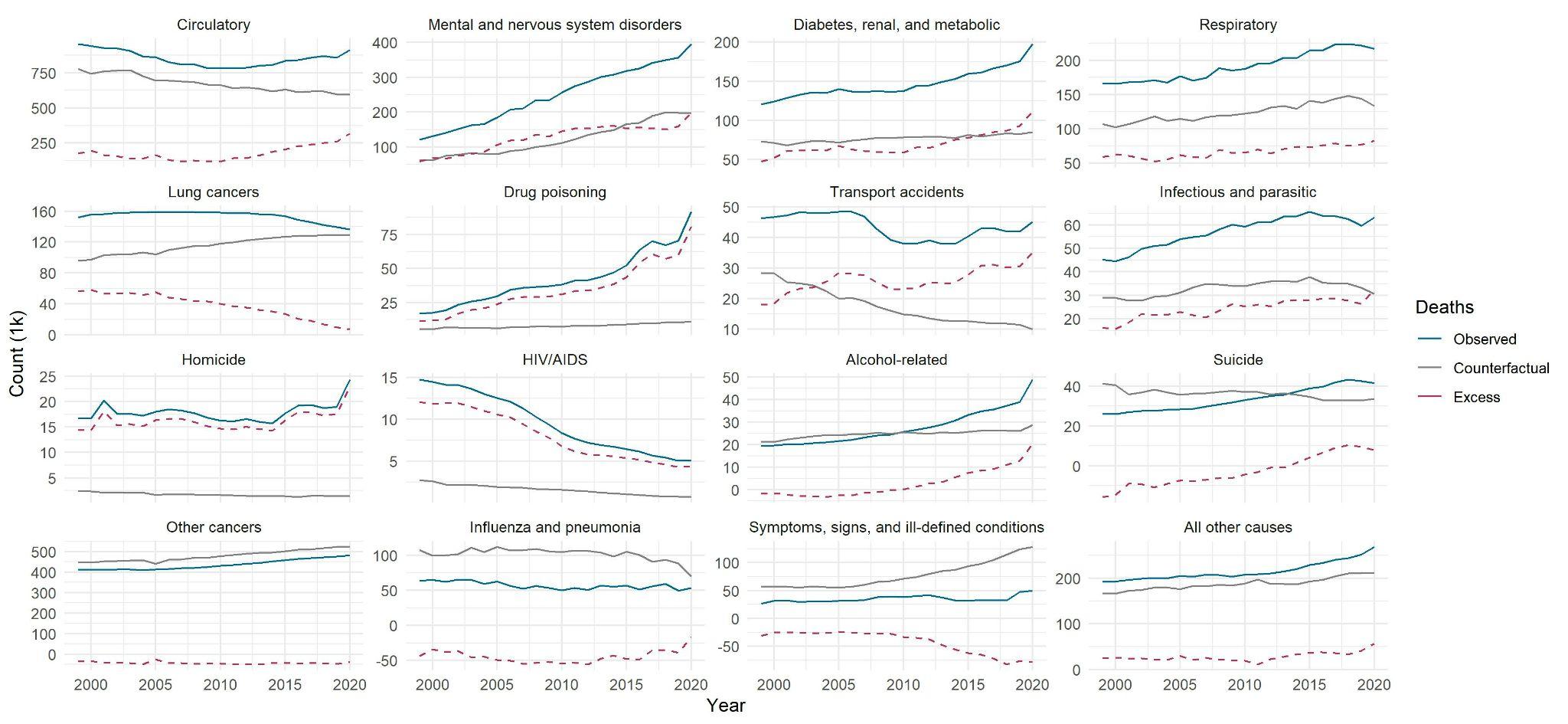

**Notes:** The counterfactual refers to the expected number of deaths the US would have experienced if it had death rates equal to the average of 12 other wealthy nations, which are described in eTable 1.

**eFigure 2.** Annual observed YLL in the US, counterfactual YLL in other wealthy nations, and excess YLL in the US compared to other wealthy nations for each cause of death from 1999 to 2020

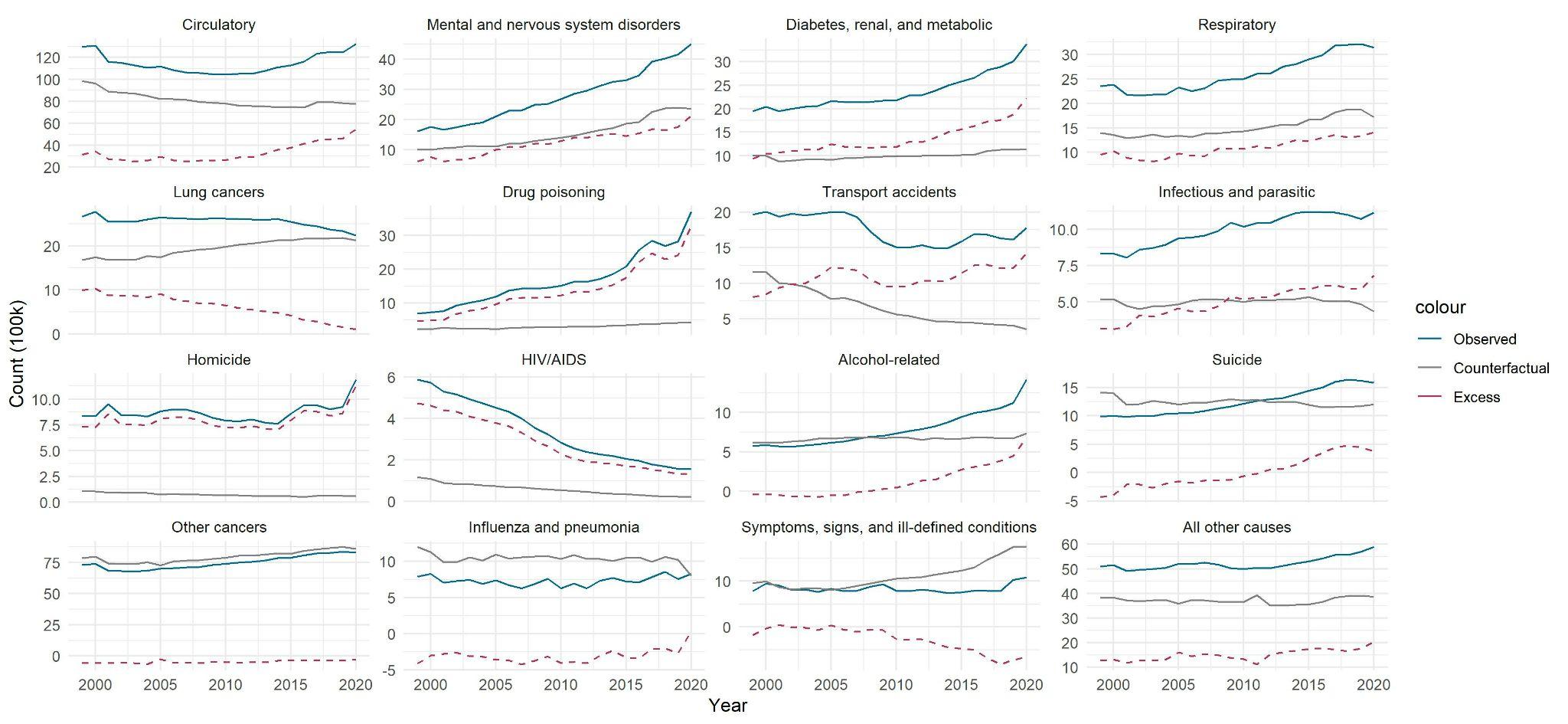

**Abbreviations:** YLL = Years of Life Lost

**Notes:** The counterfactual refers to the expected number of YLL the US would have experienced if it had death rates equal to the average of 12 other wealthy nations, which are described in eTable 1.

##### eFigure 3. Annual excess deaths by age-group in the US compared to other wealthy nations by cause of death from 1999 to 2020

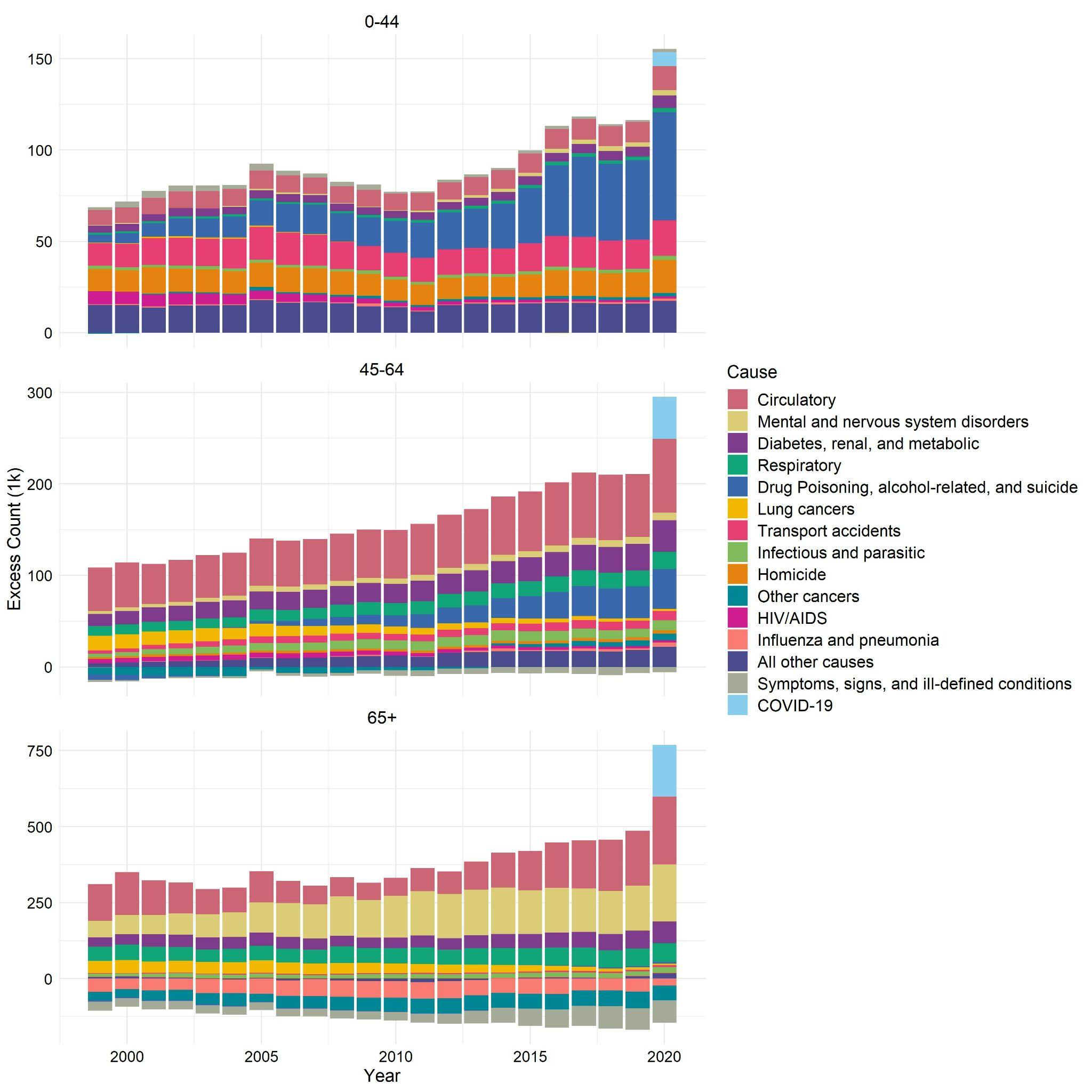

##### eFigure 4. Annual excess deaths and excess YLL by age-group in the US compared to other wealthy nations by cause of death from 1999 to 2020

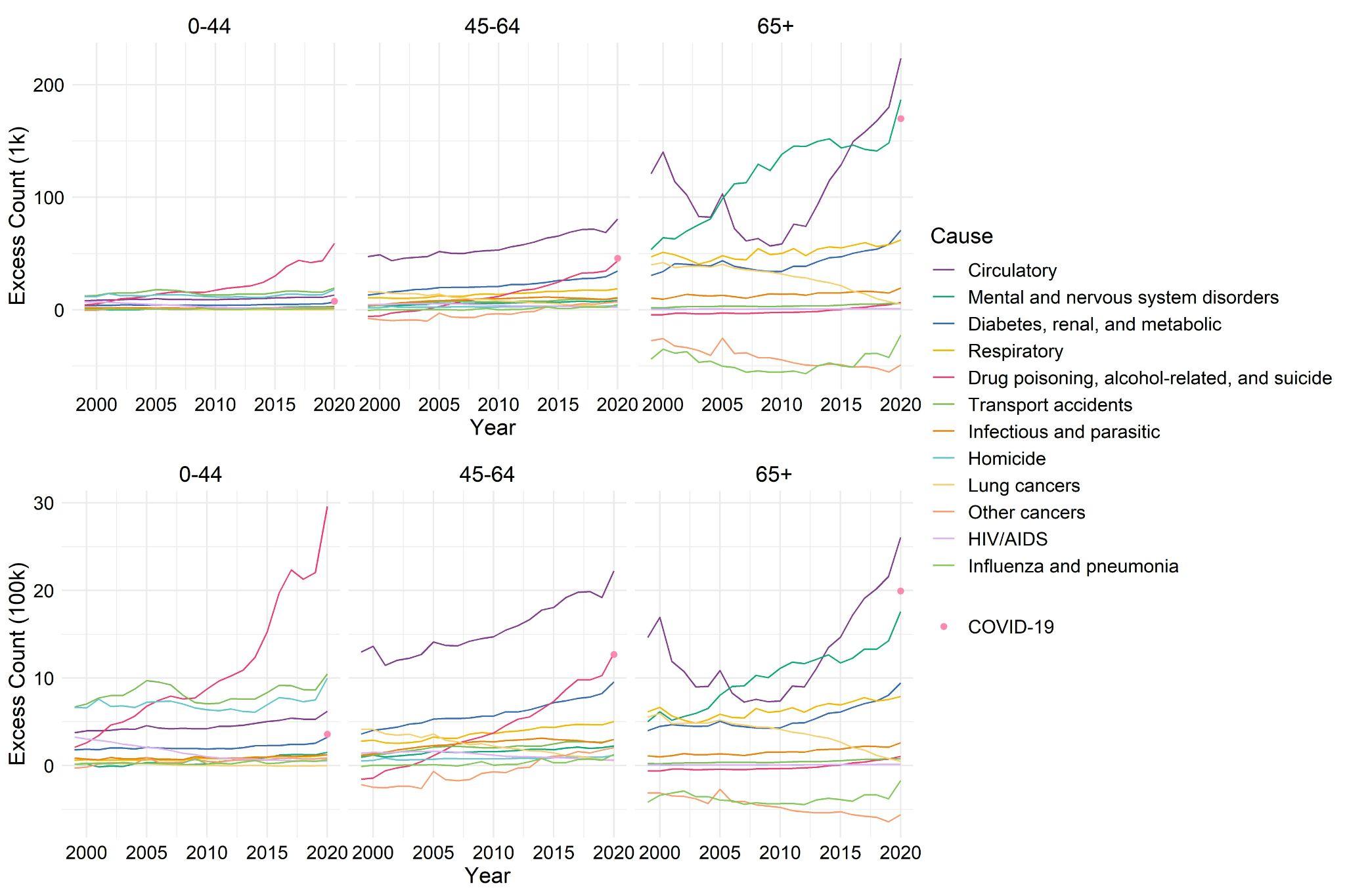

##### Supplementary Data & Replication Code

The underlying data used in the study are publicly available from the World Health Organization and the Human Mortality Database. The estimates generated in this study and programming code for replicating the analyses can be downloaded from the following permanent repository: <https://osf.io/85sk2/>
